# Does Time to Theatre Affect The Ability to Achieve Fracture Reduction in Tibial Plateau Fractures?

**DOI:** 10.1101/2021.09.27.21264218

**Authors:** David S. Kitchen, Jack Richards, Peter J. Smitham, Gerald J. Atkins, Lucian B. Solomon

## Abstract

**Aims:** The primary aim of this study was to assess the effect of time to surgery on fracture reduction, assessed as residual articular step, in cases of tibial plateau fracture (TPF). The secondary aim was to assess the effect of pre-operative demographics and residual articular step on patient reported outcomes (PROMs) following TPF.

**Method:** Between 2006 and 2017 all surgically treated TPF patients managed by a single surgeon at our institution were prospectively consented for the study of fracture outcomes. Timing to surgical intervention, reduction of articular step, age, gender, medical background, fracture classification, mechanism of injury and PROMs (Lysholm Scores and Knee Injury and Osteoarthritis Outcome Scores (KOOS)) were recorded and analysed. Reduction of articular step, defined as <2mm, was assessed by a single blinded examiner using measurements on plain radiographs on PACS.

**Results:** One hundred seventeen patients were enrolled, 52 with Schatzker II, four with Schatzker IV and 61 with Schatzker VI fractures. Patients were followed-up to a mean time of 3.9 years. The ability to achieve fracture reduction was negatively influenced by time to theatre with the odds of achieving reduction decreasing 17% each day post-injury (p = 0.002). An increased time to theatre was associated with reduced Lysholm scores at the one-year mark (p = 0.01). The ability to achieve fracture reduction did not influence PROMs within the study period.

**Conclusion:** Delay in surgical fixation negatively affects fracture reduction in TPF and may delay recovery. However, residual articular step did not influence the investigated PROMs in the cohort investigated over the mid-term (mean of 3.9 years).

## Introduction

Patients with a displaced tibial plateau fracture (TPF) routinely undergo open reduction internal fixation with the aim to restore articular surface and joint congruency. The time from injury to surgical intervention has been shown to affect the ability to reduce acetabular fractures (1, 2). This time interval might be similarly important for the reduction of other articular fractures, including TPF. The decreased ability to achieve fracture reduction with increased time from injury is likely multifactorial, with the initiation of the inflammatory phase of fracture healing likely involved. Callus formation, in conjunction with local soft tissue swelling pose challenges to achieving reduction. In addition to increased difficulty in manipulating fracture fragments as healing progresses, cancellous bone loss, occurring as early as five days post-injury in the fractured tibial plateau (3), may also contribute to impaired fracture reduction.

Adequate reduction of articular step-off was suggested to be the single biggest determinant of clinical outcomes in lower limb articular fractures (4, 5). Inadequate fracture reduction alters the force distributed onto the tibial plateau leading to an axial overload and subsequent increased rate of joint degeneration (6).

TPFs are associated with soft tissue injury (7), which is a common cause for delayed surgical intervention (8) in an effort to decrease potential surgical site infections (SSI). Whether increased time to theatre detrimentally affects the ability to reduce TPFs remains unknown. Therefore, the primary aim of this study was to assess the effect of time to surgery on the ability to reduce the articular step in TPFs. The secondary aim was to assess the effect of residual articular steps and pre-operative demographics on PROMs, specifically the Lysholm and the Knee injury and Osteoarthritis Outcome Scores (KOOS) scores, following TPF. Whether time to theatre influenced SSI in cases of TPF was also analysed.

## Methods

### Patients

This study was approved by the human research ethics committee of our institution, a tertiary referral public hospital (No. 080107/2008). A consecutive cohort of patients with unilateral TPF were prospectively consented to participate in the study between September 2006 and October 2017. All patients were managed under the care of a single surgeon (L.B.S). Patients with unilateral TPFs and an articular step-off who were treated surgically were included in the study. Patients with open injuries (5), no articular depression or fracture displacement (2) and those who declined participation in the study (28) were excluded. Follow-up review was scheduled at 6 weeks, 3 months, 6 months, 1, 2, 4, 5, 7 and 10 years, with radiographs performed at each review. Demographic data captured included patient age, gender, pre-operative smoking status and diabetic status. Patients with post-operative surgical site infections were included for radiographic parameters but excluded from analysis of their PROMs due to known influence on patient clinical course (9).

Fracture type, time from injury and mechanism of injury were recorded. Mechanism of injury was classified as either low energy, such as from a simple fall, or high energy, such as that sustained in a road traffic accident (9). Fractures were classified according to Schatzker (6).

Sub-group analysis was performed on patients whose time to theatre was up to 5 days or greater than 5 days, based on previous work showing significant loss of bone in the tibial plateau in the latter group (3).

### Radiographic Assessment

Anteroposterior (AP) and lateral radiographs performed within 48 hours of surgery and from each follow-up clinical review were assessed by a single examiner (D.S.K.) blinded to patient demographic factors and clinical outcomes. Assessment of the degree of reduction of articular step and pre-operative joint depression was performed utilising the picture archiving and communication (PACS) software at our institution. Fracture reduction was assessed on radiographs taken on the 1^st^ or 2^nd^ postoperative day. Fractures were defined as reduced if residual articular steps were less than 2mm with steps greater than 2mm being defined as not reduced (2, 10, 11). Overall alignment and condylar width were not included in the assessment of fracture reduction.

Pre-operative and post-operative joint depression was assessed using both AP and lateral radiographs. The step-off of the articular fragment with the most displacement in either radiograph was determined and analysed (Figures 1 and 2).

**Figure 1.**
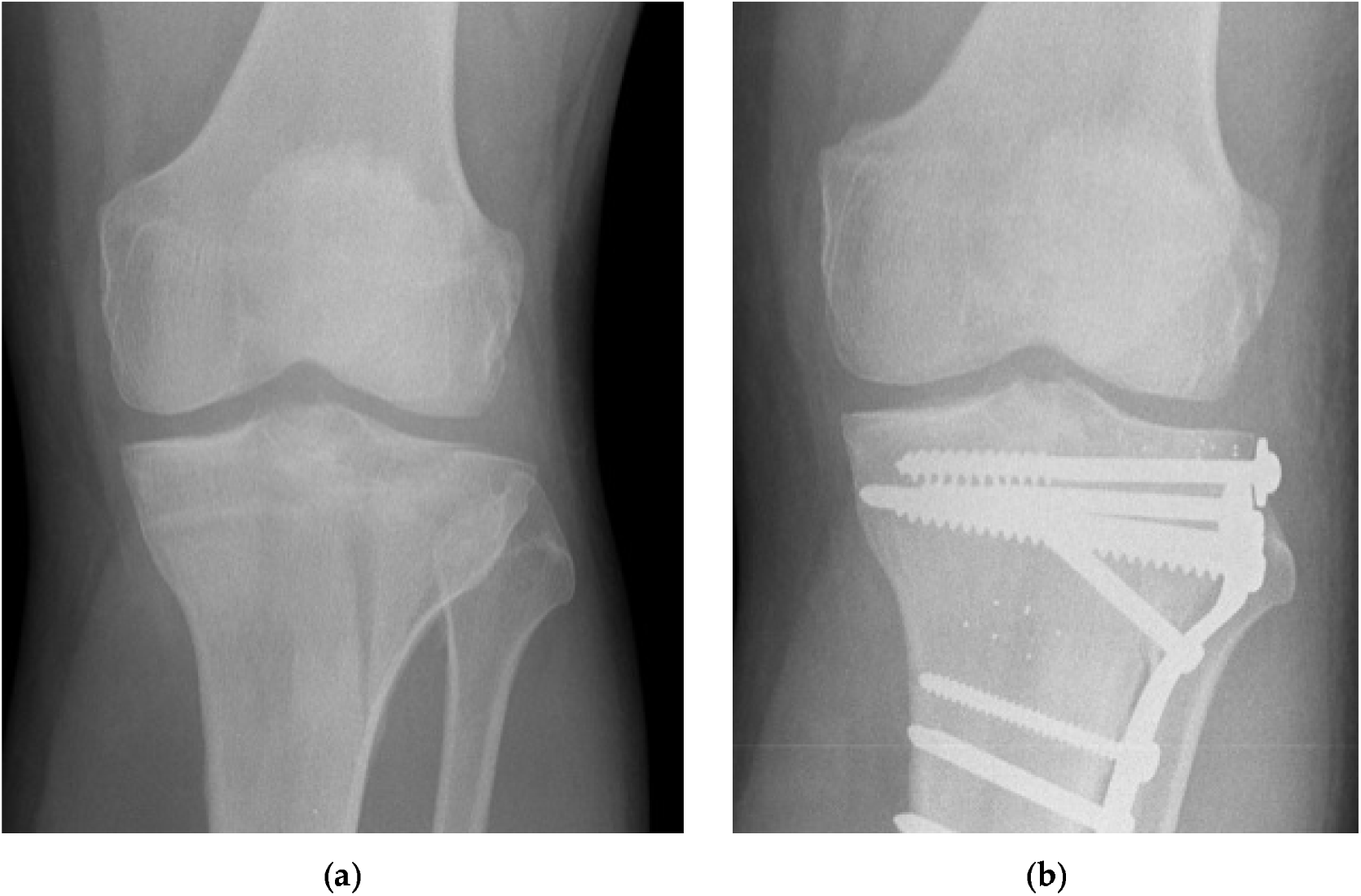
Pre (a) and postoperative day 2 (b) AP radiographs of a reduced tibial plateau fracture (Schatzker II) in a patient without medical co-morbidities.

**Figure 2.**
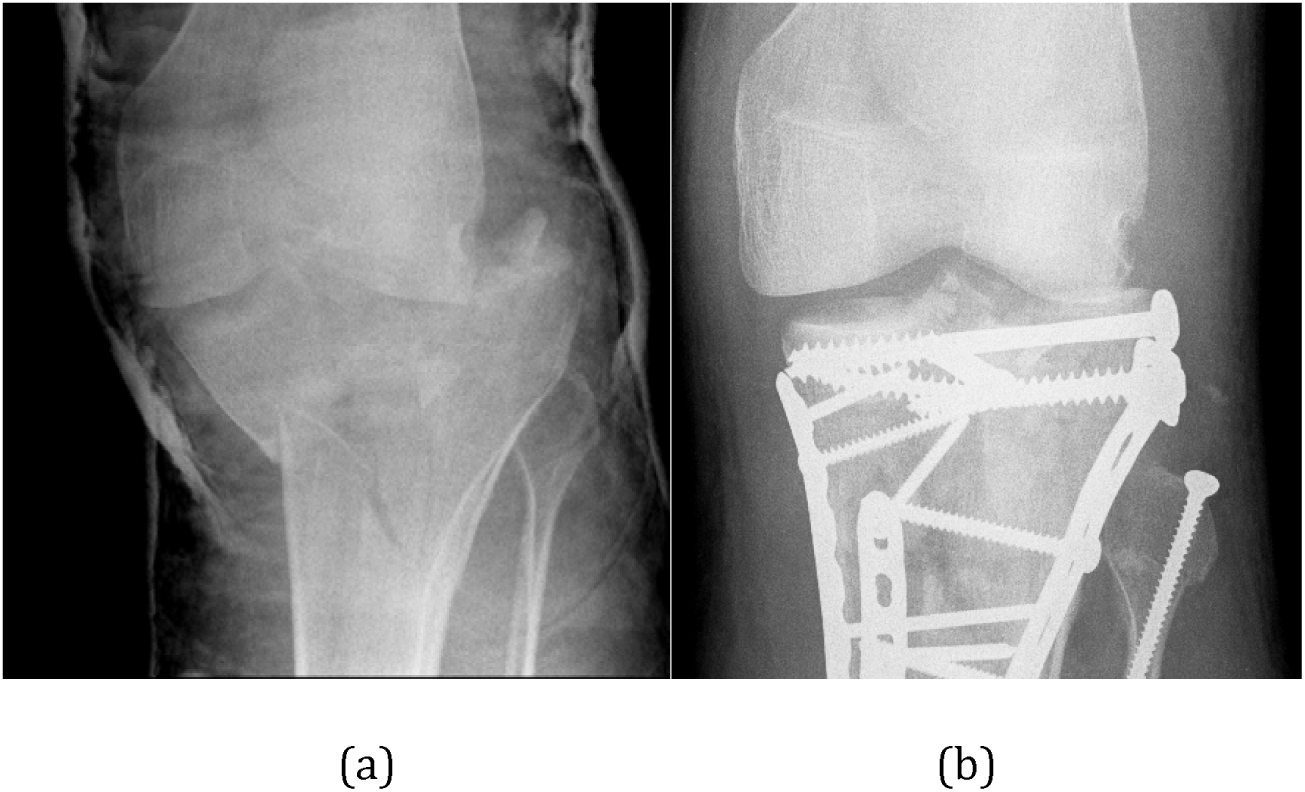
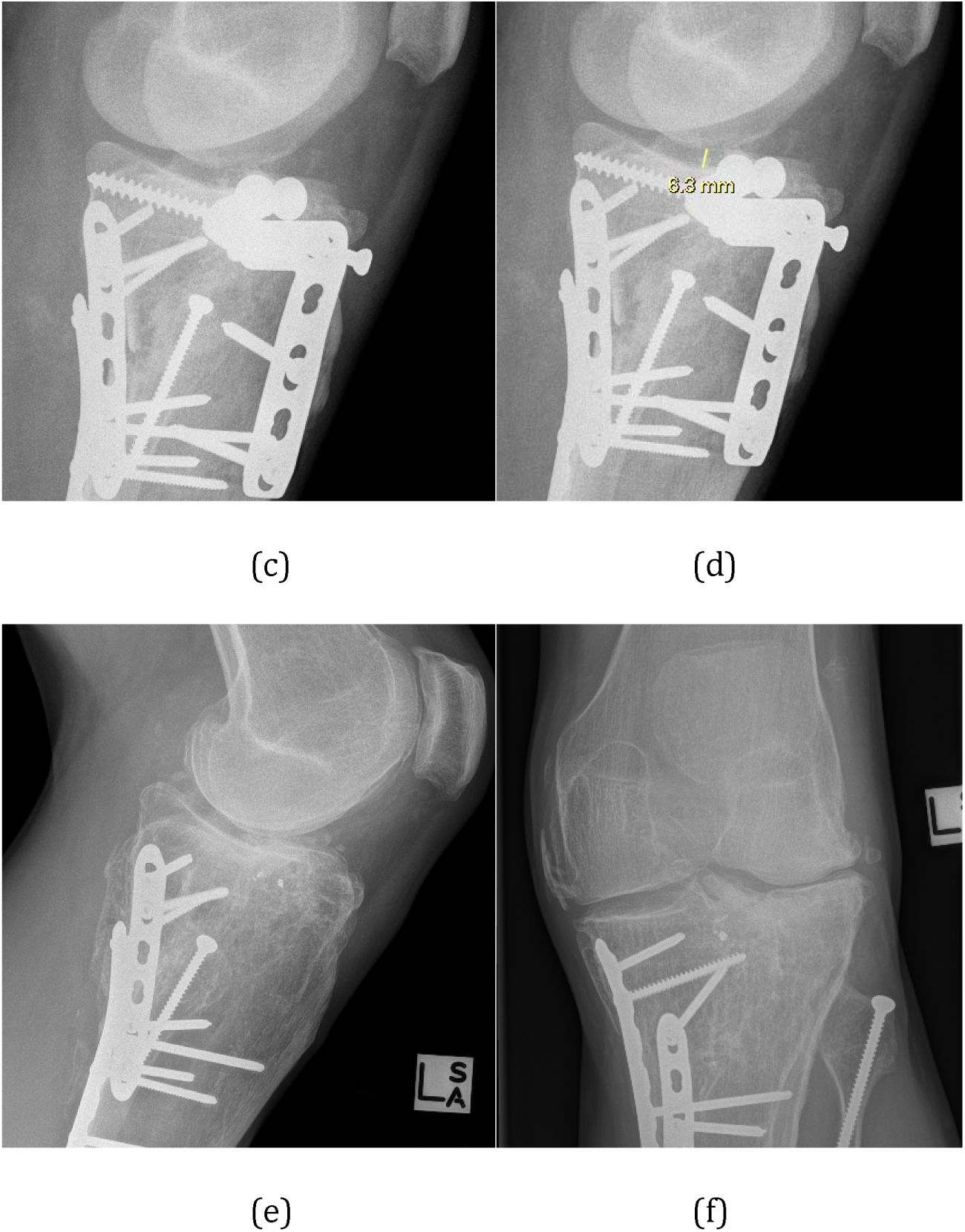
Pre (a) and postoperative day 2 (b) AP radiographs of a fracture that was not reduced (Schatzker VI) in a patient with Marfan syndrome. Note that the articular step is better visible on the lateral views (c and d). e) and f) Lateral and AP radiographs of the same fracture 5 years after injury. Note the residual articular step and secondary degenerative changes. In spite of the residual step the patient had minimal functional deficit.

### Patient Reported Outcome Measures

Questionnaires assessing the KOOS and Lysholm scores were collected at each outpatient follow-up visit. The KOOS quality of life (KOOS QOL), pain score (KOOS Pain) and Lysholm scores were analysed to determine a patient’s post-injury level of function and symptomatology. Data at 4-5 years follow-up were clustered into a ‘mid-term’ time point. If patients attended at both four and five years, then the five-year time point was utilised.

### Data Analysis

To determine the association between time to theatre and fracture reduction, both unadjusted and adjusted logistic regression models were fitted. Patient demographics, fracture type and mechanism of injury were incorporated into the adjusted model. The effect of these factors as well as fracture reduction and pre-operative step on PROMs were also analysed. This was undertaken utilising two-tailed, unpaired Student’s t-test, mixed-effect linear regression analyses and Fisher’s Exact test, with significance assumed to be for *p* ≤ 0.05. Analyses were performed using GraphPad Prism Software (version 7; GraphPad, La Jolla, CA) and Stata (version 15.1; StataCorp, College Station, TX). Mean values with ranges were measured with odds ratio and 95% confidence intervals measured for the logistic regression models.

## Results

### Patient demographics and fracture type

One hundred fifty-two eligible patients were identified with 35 patients excluded, as described above. The final cohort of 117 patients consisted of 74 males and 43 females. Demographic data including fracture type, fracture reduction and time to theatre are shown in Table 1. The overall mean time to theatre for all patients was 5.9 (0-26) days. Sixty-one patients were taken to theatre ≥ 5 days after their injury (52.1%) with the remaining fifty-six being operated < 5 days from injury (47.9%). Demographic status, including fracture type (p = 0.17), mechanism of injury (p = 0.91) and gender (p = 0.48) were found not to influence time to theatre.

**Table 1.** Patient demographics and their relationship to time to theatre.

| **Demographic** | **Sub-Category** | **Value (Range)** | **Mean time to theatre (SD, range) (days)** | **p value** |
| --- | --- | --- | --- | --- |
| Age (years) |  | 45.4 (21-78) |  | 0.388 |
| Sex | Male | 74 | 6.4 (5.0, 0-26) | 0.129 |
|  | Female | 43 | 5.1 (3.5, 1-19) |  |
| Smoking status | Smoker | 25 | 7.1 (5.1, 1-26) | 0.114 |
|  | Non-smoker | 92 | 5.6 (4.3, 0-24) |  |
| Diabetes status | Diabetic | 5 | 3.4 (1.9, 2-6) | 0.202 |
|  | Non-diabetic | 112 | 6.1 (4.6, 0-26) |  |
| Schatzker Type | I | 0 |  | 0.627 |
|  | II | 52 | 6.0 (4.7, 0-24) |  |
|  | III | 0 |  |  |
|  | IV | 4 | 3.8 (2.2, 1-6) |  |
|  | V | 0 |  |  |
|  | VI | 61 | 6.0 (4.6, 1-26) |  |
| Mechanism of injury | Low energy | 39 | 6.0 (5.1, 2-24) | 0.911 |
|  | High energy | 78 | 5.9 (4.3, 0-26) |  |
| Pre-operative articular step (mm) |  | 8.1 (0.5-54) |  | 0.009 |
| Time to theatre (days) | Overall | 5.9 (0-26) |  |  |

Patients were followed up for a mean time of 3.9 years (0.5-10). There was loss to follow up at each time point with 4% loss at 6 months, 8% at 1 year and 38% at 4 years after surgery. These losses to follow-up were due to patients failing to attend further outpatient appointments. For patients lost to follow up, on review of the reduction of their fractures 80% of those who failed to attend at 6 months had reduced fractures, 89% for those at 1 year and 73% at the mid-term. The percentage completion rate for PROMs scores at various time points was 70% at 6 months, 81% at 1 year and 93% as the mid-term time point, with all patients having follow up radiographs taken at each review.

Two patients had superficial SSI post operatively (1.7%), of these one was taken to theatre day three and the other day 10 post–injury.

### Determinants of fracture reduction

Radiographic assessment revealed that fracture reduction was achieved in 77 cases, with 40 fractures being mal-reduced. Demographic data, including pre-operative joint depression (p = 0.47) and mechanism of injury (p = 0.17) were found not to influence fracture reduction (Table 2). Time to theatre was shown to have a significant relationship with the ability to achieve fracture reduction (p = 0.002, OR = 0.83, 95% CI 0.74 to 0.93), with the odds of obtaining reduction decreasing 17% with every subsequent day post-injury (Table 2). Patients with a time to theatre of ≥ 5 days were significantly less likely to have their fracture reduced when compared with those with a time to theatre of < 5 days (p = 0.009, OR = 0.28, 95% CI 0.11 to 0.73). Patients with reduced fractures were operated on between days 0 and 24 with a mean time to theatre of 4.8 days following injury. Patients whose fractures were not reduced had been treated between days 1 and 26, with a mean time to theatre of 8.0 days. Increased pre-operative articular step was associated with an increased time to theatre (p = 0.009), but this effect was not shown when assessing patients taken to theatre < 5 days with those taken ≥ 5 days (p = 0.309).

**Table 2.** Association between time to theatre and fracture reduction

| Exposure |  | Adjusted OR<br>(95% CI) | p value |
| --- | --- | --- | --- |
| Time to theatre | Days | 0.83 (0.74, 0.93) | 0.002 |
| Age |  | 1.03 (0.99, 1.06) | 0.127 |
| Sex | Male | 0.64 (0.21, 1.98) | 0.437 |
|  | Female | Reference |  |
| Smoking status | Smoker | 0.46 (0.16, 1.39) | 0.170 |
|  | Non-smoker | Reference |  |
| Diabetes status | Diabetic | 0.64 (0.05, 7.64) | 0.722 |
|  | Non-diabetic | Reference |  |
| Schatzker Type | II | Reference | 0.168 |
|  | IV | * |  |
|  | VI | 0.48 (0.17, 1.36) |  |
| Mechanism of injury | Low energy | Reference | 0.173 |
|  | High energy | 0.48 (0.16, 1.38) |  |
| Pre-operative<br>articular step |  | 0.98 (0.92, 1.04) | 0.468 |

\*Perfectly predicts fracture reduction, that is, all patients with Schatzker type IV had reduced fractures

Comparable levels of joint depression were observed in both the < 5 and ≥ 5 days time-to-theatre cohorts, with 8.08mm (1.6-43) and 8.15mm (1.5-54) respectively (p = 0.75).

### Influence of fracture reduction and type on PROMs

Neither fracture reduction, nor pre-operative joint depression, were found to influence PROMs at any of the time points assessed (Table 3). Due to small numbers in the Schatzker IV patient cohort, the influence of fracture type and PROMs was only applied to those in the Schatzker II and VI groups. Schatzker VI fracture patients were found to have poorer Lysholm scores at the 6-month mark in comparison to Schatzker II fractures (p = 0.049, Table 4). There were no other significant interactions for outcome measures at any time point.

**Table 3.** Patient reported outcome scores in reduced and unreduced fracture patient cohorts.

|  | **Reduced** | **Unreduced** | **p value** |
| --- | --- | --- | --- |
| 6 month Lysholm | 68.1 (30-100) | 62.8 (8-92) | 0.298 |
| 6 month KOOS pain | 75.0 (41-100) | 69.8 (21-94) | 0.230 |
| 6 month KOOS QOL | 48.9 (6-100) | 41.2 (6-94) | 0.151 |
| 1 year Lysholm | 70.3 (29-100) | 66.8 (23-100) | 0.438 |
| 1 year KOOS pain | 75.2 (39-100) | 76.1 (47-100) | 0.810 |
| 1 year KOOS QOL | 51.7 (0-100) | 52.3 (19-100) | 0.918 |
| Mid-term Lysholm | 73.0 (32-100) | 72.1 (34-99) | 0.875 |
| Mid-term KOOS pain | 81.6 (47-100) | 82.7 (42-100) | 0.801 |
| Mid-term KOOS QOL | 58.0 (6-100) | 62.8 (25-100) | 0.436 |

**Table 4.** Patient reported outcomes scores vs fracture type (Schatzker classification)

|  | **Schatzker II** | **Schatzker VI** | **p value** |
| --- | --- | --- | --- |
| 6 month Lysholm | 70.7 (8-100) | 60.7 (9-92) | *0.049* |
| 6 month KOOS pain | 74.1 (21-100) | 71.8 (34-97) | 0.600 |
| 6 month KOOS QOL | 51.1 (6-100) | 41.4 (6-94) | 0.067 |
| 1 year Lysholm | 70 (23-100) | 68.6 (29-100) | 0.745 |
| 1 year KOOS pain | 75.2 (42-100) | 75.4 (39-100) | 0.947 |
| 1 year KOOS QOL | 52.3 (19-100) | 50.2 (0-100) | 0.708 |
| Mid-term Lysholm | 72.6 (34-100) | 74.3 (32-99) | 0.734 |
| Mid-term KOOS pain | 84.5 (47-100) | 75.9 (56-100) | 0.105 |
| Mid-term KOOS QOL | 60.2 (6-100) | 51.0 (19-100) | 0.314 |

### Influence of time to theatre on PROMs

Time to theatre showed significant relationships with Lysholm score at the one-year mark (*p* = 0.01) and KOOS QOL at the mid-term time point (*p* = 0.044). Lysholm score and time to theatre at one year when grouped into <5 or ≥5 days showed that the mean score was 10.34 points lower (p = 0.014, 95% CI = −18.56 to −2.13) in the ≥5 days cohort. Surprisingly, an increased time to theatre was found to have a positive influence on KOOS QOL at the mid-term (Figure 3). Sub-group analysis of the effect of KOOS QOL and prolonged time to theatre showed that the ≥5 days cohort had a mean score 14.23 higher than that in <5-day group (p = 0.009, 95% CI = 3.63 to 24.84).

**Figure 3.**
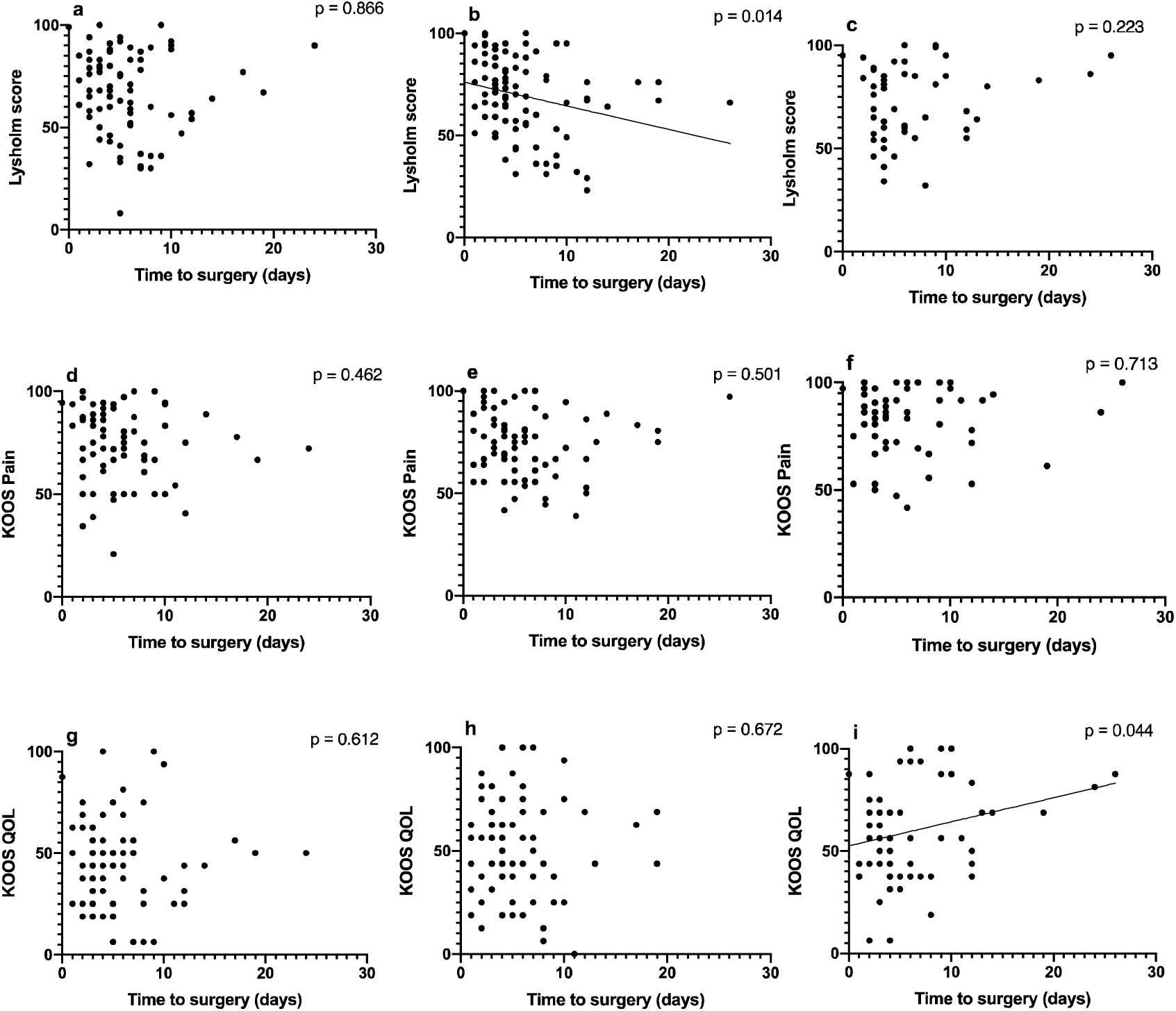
Effect of time to theatre on patient reported outcome score. Lysholm score at a) 6 months b) 12 months c) medium term. KOOS Pain at d) 6 months e) 12 months f) medium term. KOOS QOL at g) 6 months h) 12 months and i) medium term.

## Discussion

This study demonstrated a strong negative relationship between increased time to theatre and the ability to reduce TPFs. For each day of delay in fixation, the likelihood of achieving reduction decreased by 17%. A similar effect has been shown in acetabular fractures (2), with increased development of post-traumatic arthritic changes seen secondary to delays in surgical intervention (12). In our study, there was a clear difference in achieving fracture reduction when assessing patients taken to theatre on or after five days from injury, when compared to those taken before day five, with a 72% decreased chance of reduction in those taken on or after day five. Many factors can be responsible for this difference, starting with the increased difficulty in manipulating fracture fragments as fracture healing progresses. Significant cancellous bone loss around the fracture site has been shown to occur in TPFs after 5 days of injury (3). This loss of cancellous bone, may also contribute to increased difficulty in achieving fracture reduction, as fracture fragments are more susceptible to deformation during their manipulation intra-operatively.

The main reasons to postpone surgical management of TPF are soft tissue swelling, surgical time and team availability. TPFs are often associated with significant local soft tissue injury, which can compromise its blood supply and have traditionally been thought to contribute to the increased rates of post-operative SSI in cases operated within 72 hours after injury (7, 13). The effect of time to surgery on SSI in our study is inconclusive as only 2 patients (1.7%), one in each group (one operated on day 3 while the other on day 10) developed a superficial SSI. This study suggests that if the risk of SSI is not perceived as a reason for delaying surgical management of TPFs, every effort should be made to operate early, in order to provide the best chance to reduce these fractures.

To what degree articular fracture reduction influences outcomes in TPFs, is still a matter of debate. To begin with, the definition of reduction of TPF is poorly defined in the literature. The definition used in this study, that of less than 2mm articular step-off, one of the most commonly used in the more recent literature (10), does not take into consideration angular malalignment or condylar width. Although previous work linked residual articular step-off with poorer PROMs, across a range of measures including KOOS, WOMAC, the Oxford Knee score and the Musculoskeletal Function Assessment score (14, 15), we were unable to demonstrate a similar relationship between articular reduction and PROMs. Of importance, assessment of PROMs after TPF in this study was affected by patient completion, with between 70-93% of participants completing their PROMs questionnaires at each follow-up. However, other studies concur with our findings and found no correlation between articular step-off and PROMs (16-18). It is worthwhile noting that there are no PROMs specifically developed for TPF. The Lysholm score was developed for knee ligament injuries (19), while the KOOS score began as a tool to measure outcome after meniscal or ACL surgery to the knee, with its use extended to post-traumatic OA and total joint arthroplasty assessment (20, 21). Importantly, and as reported by other studies, the mean score recorded for all PROMs improved over time (22, 23). Based on the finding of improved one-year Lysholm scores in patients that were treated early, < 5 days, we can also speculate that earlier surgery, and therefore earlier commencement of rehabilitation, might lead to a faster recovery in these patients.

It is well established that patients with a more severe TPF have less favourable outcomes, with higher rates of post-traumatic arthritis and progression to total knee replacement (24). The association between the various fracture types and PROMs is less well established and there are suggestions that the TPF type does not correlate with PROMs, i.e. KOOS or Oxford Knee Scores (14). Our study tended to support this lack of interaction between fracture type and PROMs, though those with Schatzker II type injuries had higher Lysholm scores at 6 months post-operative than Schatzker VI patients. This suggests that those patients with simpler fractures may report better outcomes earlier than those with more severe injuries, however this difference may become negligible in the longer term.

This study has several limitations, including the retrospective assessment of prospectively collected data. This retrospective assessment in turn contributed to the reduced percentage completion rates for PROMs questionnaires. Participation in follow-up review remains a challenge in the trauma setting (25, 26) and those with poorer outcomes may be less incentivised to attend for follow-up review, potentially skewing the data and interpretation. However, for those patients lost to follow up in this study, the vast majority of them had reduced fractures (73-87%), suggesting that fracture failure to achieve fracture reduction is not contributory to loss to follow up.

Another study limitation may be the use of PROMs that are not specifically targeted towards the injury in question. Although, KOOS and Lysholm scores are not designed to look explicitly at outcomes after TPFs, they are both commonly used in this setting (14, 27). Additionally, the fact that articular step-off was the only measure utilised to determine fracture reduction could be seen as a limitation, especially given that knee stability have been suggested to be at least as important in TPF PROMs (range of motion, KOOS, WOMAC) (14, 28).

## Conclusion

We conclude that delay in surgical fixation negatively affects the ability to reduce articular steps in TPFs. The odds of achieving fracture reduction in TPFs decreases by 17% everyday post injury. Fracture type and mechanism of injury did not influence fracture reduction or time to theatre.

## Data Availability

The data presented in this study are available on request from the No external runfindcorresponding author.

